# Engaging Zimbabwean men and stakeholders in the co-adaptation of peer-delivered HIV self-testing: iterative prototyping of the IMPERATIVE Trial

**DOI:** 10.64898/2026.08.10.26360082

**Authors:** Maureen McGowan, Rufurwokuda Maswera, Louis Chisvo, Louisa Moorhouse, Freedom Dzamatira, Phyllis Mandizvidza, Blessing Tsenesa, Wilfred Otambo, Maxime Inghels, Guy Harling, Paul Mee, Till Bärnighausen, Simon Gregson, Constance Nyamukapa, Frank Tanser, Morten Skovdal

## Abstract

**Introduction:** HIV testing and pre-exposure prophylaxis (PrEP) are efficacious HIV prevention strategies, but uptake remains low among Sub-Saharan African men. Peer-delivered approaches may improve engagement. We developed an intervention combining peer-delivered oral HIV self-testing (HIVST) with incentivized peer referral to HIV services and an SMS-based HIV risk assessment among men in eastern Zimbabwe (IMPERATIVE Trial: NCT06370923). We co-adapted the intervention through iterative prototyping (IP) to enhance its acceptability, feasibility, and potential effectiveness.

**Methods:** From November 2023 to June 2024, we implemented a novel IP framework to refine and test the intervention. Four primary distributors (PDs) were trained to deliver HIVSTs to three peers and refer them to clinic services. Peers could become secondary distributors (SDs), obtain HIVSTs from community hubs and distribute them further. Qualitative data were collected alongside intervention testing to adapt the intervention over two iterations. Activities included three forum theatre workshops, one community advisory board meeting, 25 in-depth interviews, four focus group discussions, and eight observational reports involving men, implementers, stakeholders, and advisory board members. Additionally, 20 men completed baseline and one-week follow-up surveys. Quantitative data were analysed descriptively; qualitative data were analysed using thematic analysis.

**Results:** During testing, HIVST uptake was 100% among PDs, 90% among PD-recruited peers, and 63% among SD-recruited peers. Among self-testers, 50% sought confirmatory testing and about one-quarter initiated PrEP (PDs 25%, PD-recruited peers 30%, SD-recruited peers 25%). Participants viewed the intervention positively and anticipated increased HIV testing and PrEP initiation. Four areas for refinement were identified: recruitment, information dissemination, incentives, and socio-cultural factors. Participant recommendations were adopted before randomised controlled trial testing.

**Conclusion:** Peer-delivered HIVST with referral to HIV services shows promise for engaging Zimbabwean men. The IP framework incorporating participant recommendations enhanced intervention design and delivery within the IMPERATIVE trial. This methodology may inform future intervention development in similar settings.

**Summary Sentences:** *What is already known on this topic:*

➢ Men in Sub-Saharan Africa, including Zimbabwe, continue to be vulnerable to HIV infection and face challenges HIV testing and initiating services at clinics. A promising method to support HIV testing is through peer-delivered HIV self-testing; yet little is known about its acceptability, feasibility and potential efficacy among heterosexual men.

*What this study adds:*

➢ To enhance the peer-delivered intervention, we co-adapted the model with input from men, implementers, stakeholders, and advisory board members utilizing a novel iterative prototype testing and multi-method qualitative framework.
➢ We found that peer-delivered HIV self-testing and peer referral to HIV services may be an acceptable and feasible strategy to reach Zimbabwean men vulnerable to HIV and lead to high levels of HIV testing and pre-exposure prophylaxis uptake. Our qualitative research indicated four areas for intervention refinement (recruitment, information dissemination, incentives, and socio-cultural factors) which we adapted to enhance the intervention prior to scale-up in a randomised controlled trial.

*How this study might affect research, practice or policy:*

➢ Our study indicated that a peer-delivered HIV self-testing intervention may an acceptable method to reach heterosexual men in Sub-Saharan Africa and should be considered in future intervention practice. Our iterative prototyping approach was effective for co-adapting the intervention and should be considered as a research methodology, particularly in low-resource contexts aiming to rapidly implement HIV prevention interventions.

## 1. INTRODUCTION

Peer-delivered HIV self-testing (HIVST) is gaining traction as a potential approach to improve HIV testing uptake and service linkage among men (1). To ensure that peer-delivered HIVST is an acceptable, feasible, and effective intervention for Sub-Saharan African men, the intervention must be co-adapted with inputs from stakeholders, including men themselves.

Although Zimbabwe has made substantial progress in reducing new HIV infections over recent decades (2–5), men continue to experience high rates of HIV infection. This has been attributed to limited awareness of their own vulnerability to HIV, engagement in behaviours associated with HIV (e.g., inconsistent condom use), and prevailing masculinity norms (e.g., multiple concurrent partnerships) (6–11). Limited engagement with routine HIV testing and clinic-based services further delays initiation of HIV prevention (pre-exposure prophylaxis [PrEP]) or treatment (antiretroviral therapy [ART]). These delays are driven both by clinic-level barriers (e.g., clinic opening hours) and gender norms, including the perception that clinics are female spaces (3,6,11–14).

Peer-delivered HIV prevention services have emerged as effective strategies for engaging populations with high social connectivity who face barriers accessing clinic-based HIV services (15–18). To date, peer-delivered interventions have primarily focused on key populations (19) (e.g., men who have sex with men or female sex workers (20–23)) and more recently among adolescent girls and young women (24–26). However, less attention has been given to the acceptability, feasibility and effectiveness of peer-delivered interventions among heterosexual Sub-Saharan African adult men (16,27–29). This is despite growing evidence that men build and maintain close peer networks (30,31), are willing to receive or distribute HIVST kits to peers (28,32–34), and can support peer linkage to clinic-based HIV services (35).

Against this background, we set out to co-adapt a multi-component, male-focused intervention (to be delivered and tested as part of the IMPERATIVE trial (36) using an iterative prototyping approach. This complex intervention is a peer-delivered oral HIVST intervention enhanced with an SMS-based HIV risk assessment tool and an incentive structure to support linkage to clinic-based testing and HIV prevention or care services (i.e., PrEP or ART). Iterative prototyping has traditionally been used to develop, test, and modify specific health products, technologies, or surveys with repeated input from clients (37–41). This approach has been found to successfully identify weaknesses and improve product acceptability, feasibility, and fidelity (38,39). Considering these benefits and exploring the potential of iterative prototyping for the development of complex interventions (42); we developed a framework to inform an entire HIV intervention process rather than a specific product. Specifically, we: 1) used qualitative methods with diverse stakeholder groups to inform the intervention design, 2) tested the intervention in two iterations during which outcomes were assessed; and 3) co-adapted the intervention after each iteration based on findings from qualitative and quantitative activities.

Thus, the aims of this article are two-fold, namely: i) to describe the iterative prototyping framework developed to co-adapt a peer-delivered, multi-component intervention for subsequent evaluation in a large-scale, community-randomised controlled trial (36) and ii) to report the intervention refinements achieved through this approach. We specifically present the key areas of refinement identified by participants, the recommendations they proposed, and the adaptations made to the intervention. We hope that our multi-method iterative prototyping and testing approach will inform the development of future complex, person-centered HIV interventions.

## 2. METHODS

### 2.1. Study Setting

This study reflects the first stage of the Harnessing Male Peer Networks to Enhance Engagement with HIV Prevention Trial (“IMPERATIVE Trial”: ClinicalTrials.gov: NCT06370923) (43) ongoing in Mutasa, Makoni, Nyanga and Mutare districts of Manicaland province in eastern Zimbabwe. Iterative prototyping was specifically conducted in Samushonga Village-a rural area in Manicaland province which allowed for contextual comparability but did not contaminate the other IMPERATIVE trial sites. Manicaland province was identified as appropriate for the intervention based on the high HIV prevalence among adult men (9.8%) (44) and the local availability of existing research infrastructure provided by the Manicaland Centre for Public Health Research (MCPHR) - a collaboration between the Biomedical Research Training Institute (BRTI) and Imperial College London. MCPHR staff have more than two decades of experience building local partnerships and conducting HIV research with diverse populations in eastern Zimbabwe (4,45,46).

### 2.2. Draft IMPERATIVE Intervention

We used an iterative prototyping approach to collect feedback on and adapt a drafted peer-delivered HIVST intervention prior to testing in a community-randomized controlled trial (36). The drafted intervention was designed by combining multiple evidence-based HIV prevention strategies and engaging MCPHR research staff, a community advisory board (CAB), representatives from local implementing partners, and representatives from the Zimbabwean Ministry of Health and Child Care as has been described in detail elsewhere (36).

In short, the intervention proposed to recruit adult men (≥18 years) in eastern Zimbabwe with the support of local leaders. Following recruitment and enrolment, participants would become primary distributors and receive individual in-person training from study implementers on conducting and interpreting oral HIVST kits (through testing demonstrations), linkage to confirmatory testing at local clinics, and initiating PrEP or ART, where appropriate. Primary distributors would also be encouraged to identify three male peers who they believed could benefit from HIV testing and deliver to them HIVST kits. Each primary distributor would receive four HIVST packages: one for self-use and three for distribution. Each package would include: i) one oral HIVST kit with manufacturer instructions; ii) a study information sheet containing simplified HIVST instructions, study procedures, participating clinics, and a helpline number; and iii) a referral slip for clinic presentation to receive HIV confirmatory testing. Participants would be able to contact the helpline throughout the intervention for testing support and pre-/post-test counselling. The helpline was meant to be available from Monday to Saturday 8:00am-4:30pm. The intervention would additionally include an SMS-based HIV risk assessment tool, adapted from a national HIV risk assessment (47), allowing men to privately assess their own PrEP eligibility and to share results with healthcare workers to reduce clinic workload. Further, the intervention would be enhanced by an incentive structure in which primary distributors would receive 1 United States Dollar (USD) for confirmatory testing, $1 per delivered HIVST kit, and $1 per peer who links to confirmatory testing (maximum $7) via EcoCash, a mobile money application. After confirmatory testing and linkage to HIV services, recruited peers would have the opportunity to enrol as a secondary distributor and then visit one of two local community hubs (e.g., local shop, beerhall) to collect three HIVST kits for onward distribution; peers recruited by secondary distributors could further enrol as tertiary distributors. Secondary distributors would receive $1 per HIVST kit delivered to other peers and $1 per peer linked to confirmatory testing (maximum $6).

Healthcare workers (HCWs) and community hub operators would receive compensation for completed training (HCWs= $20; hub operators= $10.) Additionally, the owner of the community hub would receive $10 for HIVST package storage and hub operators would receive $1 per HIVST kit package successfully distributed to secondary distributors. Finally, each participating clinic would receive $60 for intervention participation in addition to 20¢ per completed data form.

The HIV tests used in this study were OraQuick HIV self-tests (OraQuick ADVANCE® Rapid HIV 1/2 Antibody Test, OraSure Technologies, Inc.), an oral fluid-based self-test with approximately 92% sensitivity, 99% specificity, and a 20-minute result time (48)

### 2.3. Iterative prototyping framework

To achieve the aims of this study, we modified an iterative prototyping framework to guide the co-adaptation of the peer-delivered, multi-component intervention. Our framework was based on the three-stage prototyping and testing approach developed by Hawkins et al. for public health intervention development (49). Specifically, Hawkins et al. implemented a consultation phase (stage one), a co-production phase (stage two), and a prototyping phase (stage three), sequentially over 18 months using multiple qualitative methods to develop drug prevention interventions in the United Kingdom prior to pilot testing (49). We retained these three design stages but implemented them concurrently rather than sequentially to reduce the time and resources required for intervention development in a low-resource context.

Our modified framework used five qualitative methods and two rounds of prototype testing to gather stakeholder feedback and refine the intervention (**Figure 1**). Forum theatre workshops were conducted in November 2023, followed by a CAB meeting in March 2024 and the first prototype testing round in April 2024, which included participant interviews and surveys. Findings were reviewed in a team workshop, and the intervention and ethics protocols were adapted as needed. A second round of data collection in May 2024 included focus groups with men, interviews with HCWs and community-hub operators, and a second prototype testing round with interviews and surveys. In June 2024, the team held a final workshop to refine the intervention before the randomized controlled trial. Observational reports and weekly meetings throughout March-May 2024 supported ongoing adaptation and safety monitoring; the full timeline is provided in **Supplement 1.**

**Figure 1.**
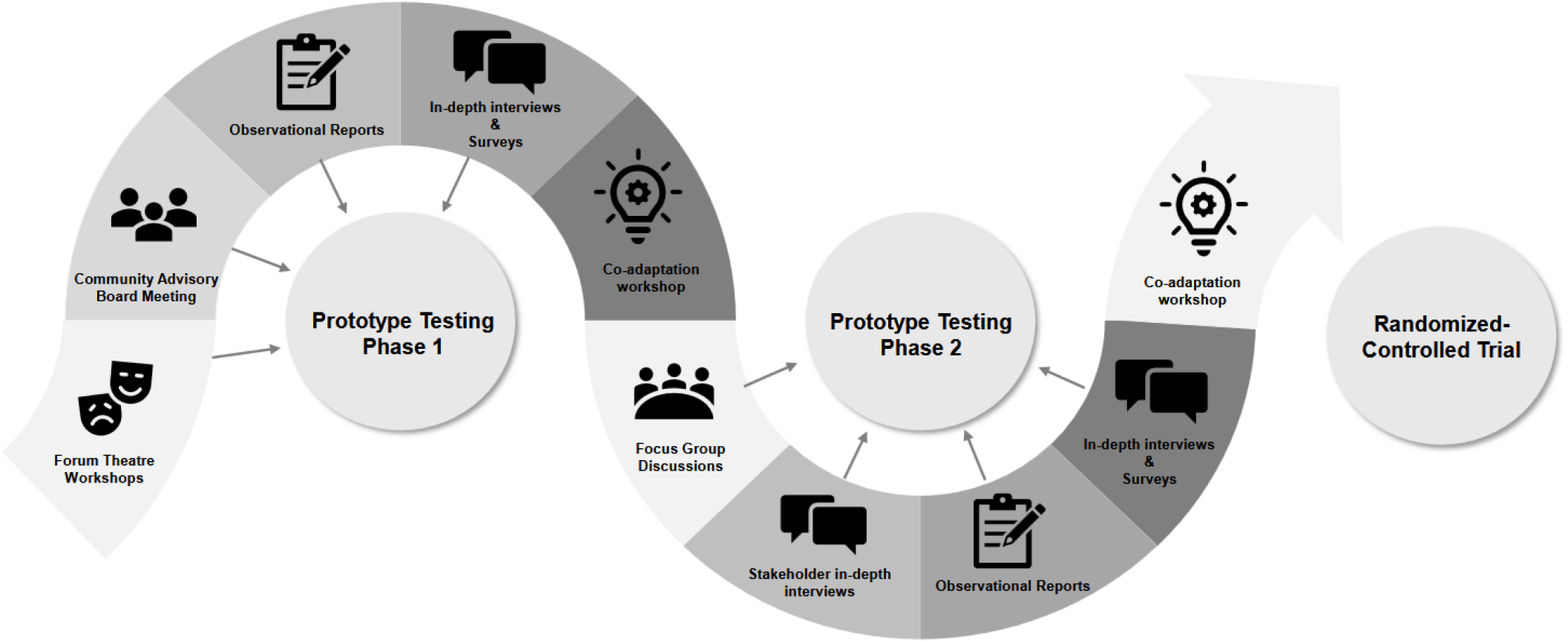
Iterative prototyping framework of a complex HIV prevention intervention.

### 2.4 Data collection

#### 2.4.1. Prototype testing

##### Recruitment

During prototype testing, four men were recruited as primary distributors to deliver the intervention to peers. Two primary distributors were recruited in the first iteration (April 2024) and two distributors were recruited in the second iteration (May 2024). Men were eligible to be primary distributors if they were ≥18 years old and resided in Samushonga Village. Preferred primary distributors were men who were HIV-negative (although this was not an eligibility criteria), men with large social networks, and those who were “respected” by and “well connected” in their communities. Prospective primary distributors were identified by village healthcare workers and village guides as well as through a sensitization meeting held by the MCPHR study implementers in Samushonga Village. Recruitment was carried out by MCPHR staff.

Participating clinics and community hubs for the iterative prototyping activity in Samushonga Village were identified and engaged to assist with the intervention and research by the study implementers. At the time of this study, the two community hubs available and willing to participate in the intervention were beerhalls.

Following identification and recruitment, the primary distributors, HCWs and community hub operators were trained to implement the intervention procedures described above in section 2.2.

##### Quantitative Procedures

Primary distributors and their peers (n=14), as well as peers recruited by secondary distributors (n=6), completed baseline surveys capturing socio-demographics, HIV testing history, service-seeking, sexual behaviours, social networks, and HIV prevention practices. Survey items were informed by the Manicaland General Population Cohort (45), a national HIV risk assessment (47), and household wealth measures developed by Schur et al (50). Seven days after enrolment, primary distributors were contacted by the study helpline team to assess intervention outcomes, including own HIVST use, kit distribution (including peer contact information), linkage to confirmatory testing, use of the SMS HIV risk assessment tool, and incentive receipt.

Peers recruited by primary distributors were contacted after seven days to complete enrolment, baseline, and outcome assessments at one time-point. They were also asked whether they had enrolled as secondary distributors, received additional HIVST kits from community hubs, and distributed them onward; corresponding quantitative data were collected from peers recruited by secondary distributors.

Baseline surveys for primary distributors were conducted in person at the MCPHR Centre, while follow-up surveys and all peer data collection were conducted via telephone by the helpline team (including author FD). Data were collected in Shona or English on tablets using Open Data Kit, and PrEP or ART initiation was verified through clinic data forms by author PM.

#### 2.4.2. Qualitative data collection

##### Recruitment

In addition to quantitative data collection, qualitative data was collected prior and in parallel to prototype testing to elicit feedback, identify areas for refinement, and collect recommendations. Qualitative data were collected among five participant groups; namely, men with or without intervention testing experience, HCWs with or without intervention testing experience, community hub operators, study implementers, and CAB members.

Individuals were eligible to participate in qualitative activities if they were ≥18 years old, male or female, and resided in Samushonga Village or any other IMPERATIVE trial sites (Mutasa District, Makoni District, Nyanga District and Mutare City) to maximize intervention acceptability, feasibility and potential effectiveness across sites.

Men without intervention testing experience were recruited either by village healthcare workers or guides and referred to the study team for consenting and enrollment procedures. HCWs and community hub operators were recruited by study implementers. Additionally, CAB members were recruited by community leaders from each of the trial study sites (2 members/study site) and referred to the study team for enrolment.

##### Qualitative Procedures

The iterative prototyping framework combined five qualitative methods: forum theatre workshops, a CAB meeting debrief, in-depth interviews (IDIs), focus group discussions (FGDs), and observational reports. Three forum theatre workshops (51,52) with men (n=12), HCWs (n=12), and study implementers (n=11) were conducted in November 2023 to assess feasibility, refine intervention components, and adapt the Theory of Change framework (23,53). Participants discussed and acted out intervention components, identified local resources to support implementation, identified implementation challenges, and proposed alternative approaches to challenges. Thereafter, a CAB meeting (including 15 CAB members) in March 2024 reviewed the cultural and contextual appropriateness of the intervention and advised on implementation, recruitment, randomization, and dissemination.

Additionally, informal IDIs with men (primary distributors=4; peers=11), HCWs (n=8), and community hub operators (n=2) explored experiences with intervention delivery and receipt, perceived barriers and facilitators, and views on HIV self-testing and PrEP services. Because the toll-free helpline number and SMS risk-assessment tool were not fully operational at the time of this study, interviews also explored anticipated perceptions of these components. Men were instead provided airtime and private phone numbers of the study implementers to compensate for the toll-free helpline number. These IDIs were supplemented by four FGDs conducted in May 2024 with men (n = 33, 8–9 men per FGD) who did or did not test the intervention. FGDs examined acceptability, feasibility, peer-network dynamics, and broader contextual influences on HIVST and PrEP uptake.

Throughout this study eight observational reports were also conducted where men and intervention stakeholders were observed carrying out intervention components to better understand how participants and stakeholders engaged with the intervention in a real-world setting.

All interview topic guides were semi-structured and designed with input from both local (RM and LC) and external (MM and MS) qualitative researchers. Each interview guide was reviewed and agreed upon via consensus by the entire qualitative team and refined between interviews, as necessary. All qualitative activities were either conducted in private rooms at the MCPHR facility (i.e., forum theatre workshops, CAB meeting, FGDs), in private rooms at clinics or community hubs, or via the phone by Zimbabwean qualitative research staff. All interviews were conducted in either in Shona or English, according to participants’ preferences. The forum theatre workshops and FGDs were recorded, translated, and transcribed verbatim. The CAB meeting, IDIs with intervention stakeholders (HCWs and community hub operators), and IDIs with intervention participants were informal and not recorded to maintain privacy due to the sensitive nature of the study. Instead, they were summarized by author RM via debriefs (n=3). Debriefs were reviewed and confirmed by author LC, a male Zimbabwean qualitative researcher and all debriefs were read and agreed upon via consensus by the entire qualitative team. Observations were conducted either from a distance (e.g., in clinic waiting rooms) or through informal conversations conducted with verbal consent. Additional methodological details are provided in **Supplement 1**.

### 2.5 Data analyses

#### Quantitative analyses

Baseline and follow-up data were analyzed descriptively via frequencies and proportions by author LM in Stata v17 (StataCorp; College Station, TX). Participant demographics and outcomes were stratified into three participant groups: primary distributors, peers recruited by primary distributors, and peers recruited by secondary distributors (**Table 1**).

**Table 1.**
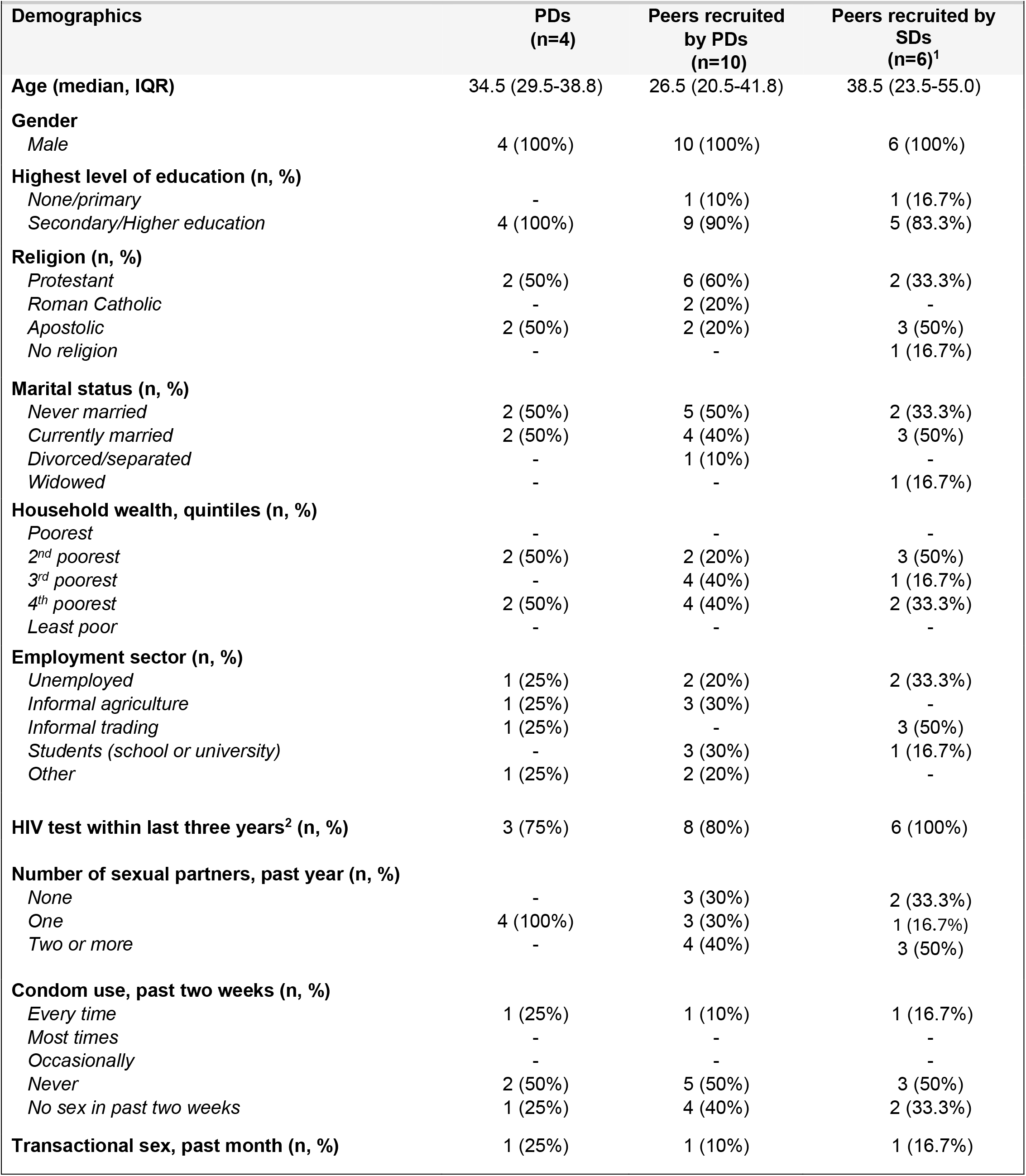
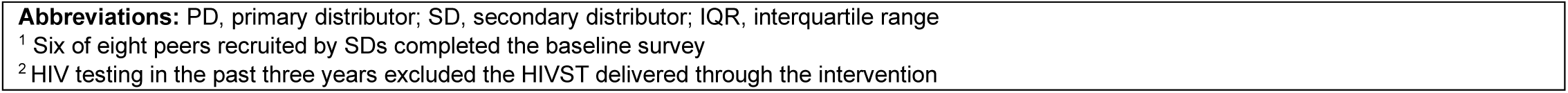
Demographic characteristics of prototype testing participants.

|  |  |  |  |
| --- | --- | --- | --- |
| Demographics | PDs<br>(n=4) | Peers recruited<br>by PDs<br>(n=10) | Peers recruited by<br>SDs<br>(n=6)<sup>1</sup> |
| **Age (median, IQR)** | 34.5 (29.5-38.8) | 26.5 (20.5-41.8) | 38.5 (23.5-55.0) |
| **Gender** |  |  |  |
| Male | 4 (100%) | 10 (100%) | 6 (100%) |
| **Highest level of education (n, %)** |  |  |  |
| None/primary | - | 1 (10%) | 1 (16.7%) |
| Secondary/Higher education | 4 (100%) | 9 (90%) | 5 (83.3%) |
| **Religion (n, %)** |  |  |  |
| Protestant | 2 (50%) | 6 (60%) | 2 (33.3%) |
| Roman Catholic | - | 2 (20%) | - |
| Apostolic | 2 (50%) | 2 (20%) | 3 (50%) |
| No religion | - | - | 1 (16.7%) |
| **Marital status (n, %)** |  |  |  |
| Never married | 2 (50%) | 5 (50%) | 2 (33.3%) |
| Currently married | 2 (50%) | 4 (40%) | 3 (50%) |
| Divorced/separated | - | 1 (10%) | - |
| Widowed | - | - | 1 (16.7%) |
| **Household wealth, quintiles (n, %)** |  |  |  |
| Poorest | - | - | - |
| 2<sup>nd</sup> poorest | 2 (50%) | 2 (20%) | 3 (50%) |
| 3<sup>rd</sup> poorest | - | 4 (40%) | 1 (16.7%) |
| 4<sup>th</sup> poorest | 2 (50%) | 4 (40%) | 2 (33.3%) |
| Least poor | - | - | - |
| **Employment sector (n, %)** |  |  |  |
| Unemployed | 1 (25%) | 2 (20%) | 2 (33.3%) |
| Informal agriculture | 1 (25%) | 3 (30%) | - |
| Informal trading | 1 (25%) | - | 3 (50%) |
| Students (school or university) | - | 3 (30%) | 1 (16.7%) |
| Other | 1 (25%) | 2 (20%) | - |
| **HIV test within last three years<sup>2</sup> (n, %)** | 3 (75%) | 8 (80%) | 6 (100%) |
| **Number of sexual partners, past year (n, %)** |  |  |  |
| None | - | 3 (30%) | 2 (33.3%) |
| One | 4 (100%) | 3 (30%) | 1 (16.7%) |
| Two or more | - | 4 (40%) | 3 (50%) |
| **Condom use, past two weeks (n, %)** |  |  |  |
| Every time | 1 (25%) | 1 (10%) | 1 (16.7%) |
| Most times | - | - | - |
| Occasionally | - | - | - |
| Never | 2 (50%) | 5 (50%) | 3 (50%) |
| No sex in past two weeks | 1 (25%) | 4 (40%) | 2 (33.3%) |
| **Transactional sex, past month (n, %)** | 1 (25%) | 1 (10%) | 1 (16.7%) |

**Abbreviations:** PD, primary distributor; SD, secondary distributor; IQR, interquartile range
<sup>1</sup> Six of eight peers recruited by SDs completed the baseline survey
<sup>2</sup> HIV testing in the past three years excluded the HIVST delivered through the intervention

#### Qualitative analyses

The qualitative data were analyzed using an inductive approach to allow participant perspectives to direct themes. In the first step, authors RM, LC, MM, and MS held team discussions to rapidly identify areas for refinement and recommendations shared by participants. These themes were labelled as potential adaptations to i) recruitment, ii) information dissemination and iii) the incentive structure. In step two, the lead author (MM) conducted a formal analysis utilizing the thematic network analysis approach by Attride-Stirling which categorized findings into a hierarchical web of basic, organizing, and global themes (54). Specifically, MM used NVivo 15 (Lumivero Version 15.3.1) to independently code the entire data corpus and arranged quotes into basic themes (e.g., peer identification, intervention awareness) which were assigned to organizing and then global themes. MM confirmed themes aligned with those identified by the study team. MM also identified a fourth area for intervention refinement, namely, to structural and socio-cultural factors. In step three, MM compared areas for refinement and recommendations to study documents (i.e., workshops documents, protocol deviations, standard operating procedures, and ethics protocol amendments) to map implemented adaptations. The final qualitative analysis was presented to the larger study team, discussed and agreed upon via consensus.

### 2.6 Public Involvement Statement

This study engaged Zimbabwean men, HCWs, community hub operators, CAB members, and study implementers. While the study team developed the initial draft intervention design and research question utilizing evidence-based research; participants were actively engaged through qualitative research methods. Additionally, the peer-delivered design of the intervention allowed for Zimbabwean men to lead peer recruitment and identify individuals whom they felt would benefit from HIV testing and prevention services. In this study, participants did not carry out analyses nor disseminate research findings. In the subsequent randomised controlled trial (36), we made adaptations to increase public involvement by including peer researchers at all stages.

### 2.7 Ethics statement

We obtained ethical clearance from BRTI (AP187/2023) and the Medical Research Council of Zimbabwe (MRCZ; A/3098). We additionally obtained ethical clearance from the University of Lincoln (UoL2024_17988), the Stellenbosch University Medical Research Ethics Committee (MREC: M23/03/009), and Heidelberg University (S-232/2026). All research activities aligned with the ethical principles stated by the Declaration of Helsinki (55).

All participants provided written informed consent (or fingerprints for those with low literacy). Participants received compensation for engagement in study activities. They received travel cost compensation (where necessary) in addition to a small remuneration which was found appropriate in the Zimbabwean context; namely, for forum theatre workshops (USD $10), FGDs and IDIs (2 bars of soap = $3), or the CAB meeting ($25).

## 3. RESULTS

### 3.1 Prototype testing

#### Participant demographics

Between April and May 2024, we successfully recruited and enrolled four primary distributors. Two primary distributors were enrolled in April 2024 in the first iteration of prototype testing, and two were enrolled in May 2024 in a second iteration of prototype testing. The primary distributors reported delivering the intervention to 10 peers in total. Three peers enrolled as secondary distributors and collected HIVST kits from community hubs and delivered kits to eight additional peers. All primary distributors and peers recruited by them completed quantitative baseline surveys and socio-demographic data. Six of the eight peers recruited by secondary distributors (75%) completed the baseline survey.

Primary distributors had a median age of 34.5 years (interquartile range [IQR] 29.5-38.8 years); peers recruited by primary distributors had a median age of 26.5 years (IQR 20.5-41.8 years) and peers recruited by secondary distributors had a median age of 38.5 years (IQR 23.5-55 years). Most participants reported completion of secondary or higher education (primary distributors 100%; peers recruited by primary distributors 90%; peers recruited by secondary distributors 83.3%). About a quarter of participants were unemployed (primary distributors 25%; peers recruited by primary distributors 20%; peers recruited by secondary distributors 33.3%) and many reported engagement in behaviours associated with HIV. Specifically, 40% of peers recruited by primary distributors and 50% of peers recruited by secondary distributors reported two or more sexual partners in the past year. Half of all participants reported never having used a condom during sex in the last two weeks and between 10-25% of men reported buying transactional sex in the past month, **Table 1**.

#### Intervention outcomes

In the first testing iteration (April 2024), both primary distributors reported using the oral HIVST kit provided to them. One attended the clinic for confirmatory testing and reported initiating PrEP. These primary distributors recruited six peers (three each), of which 83% reported using the oral HIVST kit, 50% attended confirmatory testing, and 50% reported initiating PrEP. 83% of recruited peers reported interest in enrolling as secondary distributors, but only three collected kits from community hubs and distributed these to additional peers (n=8). Of the peers recruited by secondary distributors, 63% used the HIVST kit, 50% engaged in confirmatory testing, and 25% reported initiating PrEP.

In the second testing iteration (May 2024), both primary distributors reported using HIVST kits, one attended confirmatory testing, but neither initiated PrEP. They successfully recruited four peers (two peers each). All four peers reported using oral HIVST and 50% attended a clinic for confirmatory testing. However, no peers reported initiating PrEP nor returning to the community hub to enroll as secondary distributors for onward distribution of HIVST kits.

### 3.2 Qualitative findings

All participant groups viewed the peer-delivered HIVST intervention positively. Most male participants (both those who did and did not test the intervention) described a strong interest in delivering or receiving HIVST kits through male peer networks, learning their HIV statuses, and initiating PrEP. Only a minority expressed hesitancy or mistrust being singled out and approached by peers for HIV self-testing. Overall, participants emphasized that male peers were trusted, used shared language, and were likely to maintain confidentiality of their HIV status:

> *“If we speak as men to men, it is good because we understand each other than if it’s said by women, I can just say ‘What have you seen on me?’. But if a friend says go and get tested, I will understand.” (Male participant, FGD 3)*

Although the draft intervention was generally perceived as positive; participants identified weak points requiring refinement to enhance acceptability, feasibility, and potential effectiveness. We organized these weak points into four overarching themes: i) participant identification and recruitment; ii) information dissemination; iii) incentive structure; and iv) structural and socio-cultural factors. The following sections outline the identified weak points, suggested recommendations, and adaptations made to the draft intervention.

#### Participant identification & recruitment: Both primary distributors and their peers were difficult to engage in the intervention

Foremost, both qualitative activities and prototype testing indicated that identifying and recruiting potential primary distributors and peers proved challenging (**Table 2**). At intervention onset, study implementers conducted a community sensitization meeting in Samushonga Village to raise awareness about the intervention and identify potential primary distributors; however, attendance was limited. Study implementers attributed this to gender norms, noting that women were more likely than men to attend health-related community meetings. Additionally, study implementers found that community-level HIV stigma deterred men from enrolling as primary distributors. One study implementer described:

> *“… you know the community raise their eyebrows when they see an organization interacting with a community member…. We meet at his homestead and a lot of people will start whispering around and trying to find out why he was visited by a project vehicle at his homestead. Maybe some will start spreading the rumours saying he has tested [HIV] positive and they are doing [HIV] contact tracing…” (study implementer, forum theatre workshop)*

**Table 2.** Weak-points, recommendations and adaptations to participant identification and recruitment.

| Theme | Intervention weak points | Participant recommendations that were not accommodated<sup>1</sup> | Participant recommendations that were accommodated<sup>1</sup> | Intervention Adaptations |
| --- | --- | --- | --- | --- |
| **Participant identification & recruitment** | Challenges recruiting effective PDs |  | Recruit PDs who are influential, well-liked, and trusted in the community<br>Add sensitization meetings at community-level to raise awareness about the intervention and to recruit potential PDs | Clarified PD eligibility criteria to include men living with or without HIV<br>Definition for PDs adapted from "respected" to "well liked & trusted"<br>†Included additional sensitisation meetings |
|  | Challenges recruiting peers | PDs should be identifiable to potential peers (e.g., wear branded T-shirts)<br>PDs should receive airtime to recruit peers virtually |  | *No adaptations made* |
|  | Diverse understanding of who constitutes a "peer" |  | PDs and SDs should call the helpline to confirm peer eligibility | Study implementers confirmed eligibility of recruited peers over the helpline |
|  | Intervention limited distribution to three peers each | HIVSTs should be delivered to boys and youth (<18 years)<br>HIVSTs should be delivered to sexual partners | PDs should be able to deliver kits to more than three peers | † PDs and SDs could collect a second set of three HIVST kits from community hubs for distribution |

Abbreviations: PD, primary distributor; HIVST, HIV self-test; SD, secondary distributor
<sup>1</sup> Recommendations stem from all five participant groups.
† These adaptations informed by iterative prototyping but were implemented in subsequent randomised controlled trial

Recruitment challenges were amplified by ambiguity surrounding primary distributor eligibility criteria and desired characteristics among study implementers and village guides responsible for recruitment. The intervention aimed to prioritize primary distributors with large social networks, strong community standing, and an HIV-negative status (i.e., eligible to self-test). However, village leaders interpreted “respect” in varying ways, with some associating it primarily with religious leadership. Questions also arose regarding whether men living with HIV could effectively serve as primary distributors and deliver HIVST kits to their peers, despite being ineligible to use kits themselves to prevent false-negative results.

In response, discussions were held with study implementers, HCWs, CAB members, and men to collect recommendations and co-adapt intervention recruitment strategies. A common recommendation made by participants was to provide additional sensitization meetings in communities to raise awareness about the intervention and recruit more primary distributors. Although additional sensitisation meetings were not feasible during this iterative prototyping study due to resource constraints; sensitisation was identified as a priority for future implementation and was adopted in the subsequent randomised controlled trial. In this study however, we adapted the primary distributor eligibility criteria to maximize peer network reach. For example, it was clarified for study implementers and village guides that both men living with or without HIV, were eligible to serve as primary distributors. Primary distributors known to be living with HIV (regardless of ART adherence level) would receive three instead of four HIVST kits to deliver kits to peers without testing oneself to prevent a false-positive result. Additionally, the term “respect” was replaced with locally appropriate terms such as “well-liked and trusted” to identify potential primary distributors beyond religious leaders and to encourage heterogeneity to reach diverse male social networks. A study implementer summarised desired primary distributor characteristics as follows:

> *“…sociable, friendly, trustworthy, not shy, not fearing (risk taker) and be able to approach people and articulate in a persuasive manner the purpose of the study… the person should be organized, influential and well known in the community or village for always mixing and mingling with people.*” *(Study implementer, debrief phase two)*.

Despite refinement of primary distributor eligibility criteria, they reported difficulties identifying, approaching, and recruiting their male peers. Understandings of who a “peer” was varied, with primary distributors at times recruiting relatives, neighbours, or men they perceived as likely to accept an intervention. A study implementer described peer recruitment as follows:

> *“The PDs’ [primary distributor’s] and SDs’ [secondary distributor’s] recruitment of peers was done haphazardly. Others recruited their neighbours while some recruited relatives only. However, other PDs and SDs just moved from one homestead to another trying to find whoever was 18 years, male, living in Samushonga so that they could persuade them to participate in the intervention…” (study implementer, debrief phase two)*.

Primary distributors reported that certain social groups, including football clubs, snooker (“pool”) and darts clubs, and beer-drinking groups were (or would be) easier to engage in the intervention than others. Groups including older men, men in certain occupations (e.g., farmers), men in burial societies, and men with strong religious beliefs were perceived as more difficult or culturally inappropriate to approach through a peer-delivered intervention. Male participants and HCWs noted that men from specific religious denominations would be more difficult to recruit because they may be perceived by their communities as engaging in promiscuous sexual behaviour and utilizing biomedical services over prayer and traditional healing. One male participant explained:

> *“…many [members of the Apostolic sect] will be saying that we do not go to the hospital. That is another problem that is there.” (male participant, FGD 1)*.

Conversely, some primary and secondary distributors successfully recruited peers who they felt could benefit from HIV testing and reported unmet demand within their social networks and expressed a desire to expand their reach. Some participants advocated for expanding peer eligibility to boys and youth (<18 years). Unexpectedly, a few men and HCWs also suggested distributing HIVST kits through men’s sexual networks:

> *“*…*if a person would want to include the partner, that would be good. Let’s say the result of the wife is also [HIV-] reactive, this means two people will then be assisted at the same time…” (HCW, forum theatre workshop)*

In response, participant groups shared recommendations, some of which were co-adapted into the intervention. For example, primary and secondary distributors were requested to consult the study helpline to confirm peer eligibility prior to HIVST distribution. Additionally, in the subsequent randomised controlled trial, primary and secondary distributors would be permitted to collect an additional three HIVST kit from community hubs to increase peer network reach and meet testing demand, where necessary. We did not expand HIVST delivery to youth or sexual partners as this went beyond the aim to develop an intervention specific to adult men, however, this recommendation was considered vital for future intervention models.

#### Information dissemination: Men and intervention stakeholders expressed greater need for information about the intervention, HIVST, and PrEP

A second major theme we identified was how information surrounding the intervention and HIVST and PrEP was disseminated to men, HCWs, and community hub operators (**Table 3**). Participants (particularly men and CAB members) noted that men in Samushonga Village and surrounding communities were often unaware of the intervention taking place and had limited prior knowledge or misconceptions about oral HIVST and PrEP. Although men often acknowledged PrEP’s preventative benefits; uncertainty about how PrEP worked gave them pause to engage with the intervention:

> *“Some of the things that can hinder [using PrEP] is lack of understanding how the tablets work. Some people may not understand and say that people are lying, things like this do not work.” (male participant, FGD 1)*

**Table 3.** Weak-points, recommendations and adaptations to information dissemination.

| Theme | Intervention weak points | Participant recommendations that were not accommodated<sup>1</sup> | Participant recommendations that were accommodated<sup>1</sup> | Intervention Adaptations<sup>3,4</sup> |
| --- | --- | --- | --- | --- |
| Information dissemination | Limited awareness in communities about the intervention, and HIVST or PrEP | Implement community education campaigns on HIVST and PrEP through multi-media platforms (e.g., posters, phone messages) or in male-centred locations (e.g., football fields).<br>HIVST use should be encouraged by traditional and religious leaders |  | *No adaptations made* |
|  | Participants did not read HIVST instructions or found them difficult to read | Provide HIVST demonstration videos | Adapt the study information sheet<br>Adapt HIVST packaging | † HIVST information sheet adapted to include pictorial information<sup>2</sup><br>Note and sticker added to HIVST kit to remind participants to read instructions first before use<br>HIVSTs and information sheets placed in khaki envelopes with plastic protective sheeting |
|  | Participants had limited awareness of the helpline |  |  | PDs were contacted to remind their peers of helpline availability.<br>† Adapted HIVST information sheet highlighted helpline number<sup>2</sup> |
|  | Some participants had limited confidence to HIVST | PDs should be provided with an HIVST demonstration kit | Men should be provided HIVST assistance | PDs contacted to remind their peers to call the helpline for testing support<br>(Unintended adaptation) Men sought HIVST assistance from relatives, sexual partners, study implementers, or healthcare workers |
|  | Some stakeholders had uncertainty about their intervention role | Implement train-the-trainer approaches between HCWs | Retrain HCWs and community hub operators | Implemented an SMS system for community hub operators to communicate with study database to confirm secondary distributor's eligibility prior to HIVST distribution.<br>Retrained HCWs and community hub operators<br>Developed flow chart in clinics detailing HCW responsibilities<br>† Developed WhatsApp support group for HCWs |

**Abbreviations:** PD, primary distributor; HIVST, HIV self-test; SD, secondary distributor; HCW, healthcare worker
<sup>1</sup> Recommendations stem from all five participant groups.
<sup>2</sup> The adapted HIVST instruction sheet is accessible in Supplement 1
† These adaptations were informed by iterative prototyping but were implemented in subsequent randomised controlled trial

Similarly, while HIVST was valued for its privacy, confidentiality and convenience; men expressed limited confidence self-testing, feared conducting or interpreting HIV test results incorrectly, and questioned how to receive post-test counselling. This mistrust toward HIVST may have been magnified by the intervention’s use of oral HIVST kits, as most men had not previously tested orally and questioned whether saliva could test for HIV. One participant described seeking blood-based confirmatory testing to validate the efficacy of the oral HIVST:

> *“I realised that the BRTI [Biomedical Research and Training Institute HIVST] kits were genuine and not fake when I had done the HIV confirmatory where blood was taken from me and I received the same result. I did not believe that saliva can be used to determine one’s HIV status”. (male participant, debrief phase two)*.

Despite these hesitations, most men reported using the HIVST kit delivered to them. During the testing process, most men reported that they did not read the manufacturer inserts or study information sheets – due to the length of the documents and limited time capacity- and were therefore unaware of helpline testing support and the SMS-based HIV risk assessment tool available to them. Instead, men relied on verbal instruction and testing demonstrations from study implementers or distributors. At times men also sought testing assistance from their peers, sexual partners, relatives, or HCWs, although unintended by the intervention design:

> *“Yes, hah, it [HIVST] was easy for me, and I was happy with it, because I used the kit I was given in the presence of my family, with my wife, and we did together. Hah it went well, it was exciting. And it does not take long, just a few minutes and you will be done.” (male participant, FGD 1)*.

In addition to information sought by male participants, HCWs and community hub operators also reported uncertainty regarding their roles in the intervention. For example:

> *“…health workers were not aware if they were supposed to ask participants whether they had [SMS] self-assessed themselves for HIV before coming for an HIV confirmatory test.” (study implementer, stakeholder debrief)*

Hub operators were also uncertain about how to confirm the eligibility of secondary distributors before providing them with HIVST kit packages prior to distribution.

To address these challenges, we collected recommendations from all participant groups. Expanded community education on HIVST and PrEP through multi-media platforms (e.g., posters) or at male-centred locations (e.g., football fields) was widely recommended by men and HCWs for future intervention implementation. Second, CAB members recommended to adapt HIVST packaging to include a note and a sticker saying, *“Read Me First” (CAB meeting report)* to draw attention to the manufacturer inserts and study information sheet prior to testing. In addition, we modified the study information sheet by revising and simplifying the Shona and English text, highlighted the available helpline and SMS HIV risk assessment tools, and included pictorial instructions to accommodate limited literacy among some men (**Supplement 2**). We also changed the HIVST packaging from a box to a khaki envelop sleeve with plastic protective sheeting based on feedback from study implementers and CAB members to reduce packaging costs while increasing the protection of written materials from severe weather.

Furthermore, the study team utilised feedback from prototype testing to design and implement an SMS system for community hub operators to contact the study team database to confirm the identities of secondary distributors prior to HIVST kit distribution. The study implementers retrained hub operators to utilise the SMS system and to clarify their role in the intervention. Similarly, study implementers also retrained HCWs (including newly employed staff) to clarify intervention procedures and implemented a flow chart in clinics to detail intervention responsibilities. In the subsequent randomised controlled trial, a WhatsApp group was also implemented for HCWs to enable direct communication with study implementers to address intervention queries.

#### Incentive structure: Participants discussed appropriate intervention compensation

Most participants agreed that incorporating an incentive structure would (or did) enhance motivation and effectiveness to engage with the intervention. There were, however, differing views regarding who should receive incentives and the appropriate amount (**Table 4**). In the draft intervention, only primary distributors received financial compensation, up to $7 USD ($1 for self-testing, $3 for HIVST distribution; and $3 for peers linking to confirmatory testing). Participants (including men, HCWs, CAB members, and study implementers) shared concerns that linking primary distributor’s incentives to peer outcomes could coerce peers to engage in confirmatory testing. A male participant said:

> *“Eeh, from what I am seeing, the area where problems can arise is if people will use this as a way of making money…But it also encourages. There is need to understand on the issue of the money that is given, it’s a way of encouraging and motivating each other. It’s for motivation but it can mislead others.” (Male participant, forum theatre workshop)*

**Table 4.** Weak-points, recommendations and adaptations to the incentive structure.

| Theme | Intervention weak points | Participant recommendations that were not accommodated<sup>1</sup> | Participant recommendations that were accommodated<sup>1</sup> | Intervention Adaptations<sup>3,4</sup> |
| --- | --- | --- | --- | --- |
| **Incentive structure** | Incentives may lead to coercion and corruption |  | ▪ Diversify incentives between PDs and peers to reduce risk of coercion<br>▪ Confirm SD incentive eligibility prior to incentive distribution and track incentives<br>▪ Clarify and increase transparency about incentive eligibility and procedures | 1. † Incentives were diversified; PDs incentives for self-testing and HIVST delivery. Peer incentive for confirmatory testing.<br>2. Implemented an SMS system for community hub operators to communicate with study database to determine incentive eligibility and track incentive distribution<br>3. † HIVST study information sheet clarified incentive eligibility and procedures<sup>2</sup> |
| | Incentives considered too low by PDs and peers | | ▪ Increase incentives for PDs<br>▪ Diversify incentives between PDs and peers to increase compensation for peers | 1. † Incentives were diversified; PDs incentives for self-testing and HIVST delivery. Peer incentive for confirmatory testing.<br>2. † PDs and SDs could retrieve three additional HIVST kits from community hubs and receive incentives for delivery (\$1 per HIVST) |
|  | Challenges with incentive access (e.g., delays) | ▪ Provide cash, airtime or other alternative incentives to participants | ▪ Synchronise incentive payment intervals | 1. † Synchronised incentive payments to twice weekly |
|  | Compensation was considered too low by HCWs and community hub operators |  | ▪ Increase compensation | 1. Increased compensation for data collection activities among HCWs from 20¢ to 50¢ per completed form |

**Abbreviations:** PD, primary distributor; HIVST, HIV self-test; SD, secondary distributor; HCW, healthcare worker
<sup>1</sup> Recommendations stem from all five participant groups.
<sup>2</sup> The adapted HIVST instruction sheet is accessible in Supplement 1
† These adaptations were informed by iterative prototyping but were implemented in the subsequent randomised controlled trial

Conversely, a concern raised by several men and by intervention stakeholders was that they felt incentives were generally too low. Men felt that the incentives did not sufficiently cover transportation or clinic costs. Stakeholders (HCWs and hub operators) also felt that their compensation did not appropriately reflect the work they provided. One male participant said:

> … *if you give this person a dollar to go to the clinic, he will not go, because for me to the clinic, my transport fare is $1, $0.50 to and $0.50 from, so he will not go.” (male participant, forum theatre workshop)*

Additional challenges included difficulties accessing incentive payments via the mobile money application (i.e., EcoCash). Some participants experienced long distances and transport costs to access EcoCash outlets, transaction fees, payment delays, or registration challenges:

> *“…the dynamics of using Ecocash in that, one can be send little money e.g. USD1.00 which hardly can buy something meaningful to being short changed when you want to receive hard cash as well as the fact that most EcoCash outlets’ proximity to participants is also a challenge”. (study implementer, debrief phase one)*

In response, several adaptations were made to the incentive structure prior to the randomised controlled trial. Incentives would be diversified between primary distributors and their peers, reducing coercion risk and compensating all participants appropriately. Primary distributors would receive incentives for their own HIV self-testing and HIVST kit delivery to peers. Recruited peers would receive $1 for confirmatory testing and $1 per HIVST kit distributed as a secondary distributor. In the full trial, both primary and secondary distributors would also be able to earn additional incentives if they obtained a second set of HIVST kits from the community hub and successfully distributed these.

To further reduce the risk of coercion, the SMS system developed and implemented at community hubs was also used to determine incentive eligibility and track incentive distribution. To ensure timely and transparent payments, we also synchronised incentive payments via a virtual calendar to distribute incentives twice weekly. However, increased compensation for stakeholders was not adapted to reflect real-world financial constraints. In the randomised controlled trial however, HCWs received an increase from 20¢ to 50¢ per data form to better compensate data collection activities.

#### Structural and sociocultural factors: Participants identified logistics, policies and socio-cultural norms as intervention hinderances

Participants identified structural and socio-cultural factors that they felt barred (or could bar) men’s engagement in the peer-delivered HIVST intervention (**Table 5**). For example, some participants had little engagement with the helpline, which they attributed to restricted operating hours (Monday through Saturday 8:00am-4:30pm) coinciding with their work or school hours. Study implementers also observed that men were generally challenged to engage with virtual intervention components (as well as peer enrolment and study data collection) due to limited access to phones, airtime, and/or network connectivity.

**Table 5.** Intervention weak points, participant recommendations, and implemented adaptations to structural and socio-cultural factors.

| Theme | Intervention weak points | Participant recommendations that were not accommodated<sup>1</sup> | Participant recommendations that were accommodated<sup>1</sup> | Intervention Adaptations |
| --- | --- | --- | --- | --- |
| **Structural and social-cultural factors** | Busy schedules and limited operating hours<br>limited helpline use |  | ▪ Expand helpline operating hours | 1. Helpline operating hours expanded to Monday-Saturday 8am-7pm and Sunday 8am-3pm |
|  | Far distances, busy schedules, and clinic opening hours limited<br>confirmatory testing and HIV service uptake | ▪ Conduct HIV confirmatory testing in communities<br>▪ Initiate and deliver PrEP in the community |  | *No adaptations made* |
|  | Limited phones, airtime and network connectivity challenged intervention delivery and data collection |  |  | *No adaptations made* |
|  | MoH policies and guidelines contrasted with testing procedures |  |  | 1. Retrained HCWs on confirmatory testing procedures |
|  | Gender of stakeholders could influence intervention engagement |  | ▪ Participants should have choice of male or female stakeholders throughout the intervention | 1. Male and female helpline operators made available |
|  | Beerhalls were not always considered appropriate community hub locations |  | ▪ Community hub locations should be diversified | 1. † Community hub locations diversified between beerhalls and various local shops |

**Abbreviations:** PD, primary distributor; HIVST, HIV self-test; SD, secondary distributor; HCW, healthcare worker; MOH; Ministry of Health
<sup>1</sup> Recommendations stem from all five participant groups.
<sup>2</sup> The adapted HIVST instruction sheet is accessible in Supplement 1
† These adaptations were informed by findings through iterative prototyping but were implemented in subsequent randomised control trial

When participants were asked why they may (or did) have challenges attending clinics for confirmatory testing and initiating HIV services, men often noted long travel distances to clinics, competing work and school commitments, and limited clinic opening hours as major deterrents. In addition, uncertainty about whom to provide HIV confirmatory testing to at the clinic was a concern raised by some HCWs. They reported hesitation providing confirmatory testing to men who had received an HIV-negative HIVST result, as this conflicted with previous Ministry of Health guidelines:

> *“He [healthcare worker] reported that he almost sent away a BRTI [Biomedical Research and Training Institute] participant who had come for an HIV confirmatory test as he said that the MoHCC [Ministry of Health and Child Care] testing algorithm does not allow someone who has tested HIV-negative to be done an HIV confirmatory test, hence this was a surprise to him.” (Study implementer, debrief phase two)*

One male participant also raised the concern that he perceived men to be less prioritised to initiate PrEP at clinics as compared to other populations:

> *… at the clinic, if you get there, they can say the PrEP that we have is limited, so we cannot just give anyone. So, they give to couples who have one partner who is HIV-positive and the other is negative [sero-discordant]…So, [PrEP] they are not easy to get for everyone.” (Male participant, FGD 4*)

Beyond logistic and policy constraints, socio-cultural gender norms were believed to influence intervention engagement. Nearly all men liked that HIVSTs were delivered by male peers rather than women due to fears of HIV status disclosure or stigma. A few participants (including some men, HCWs, and study implementers) similarly described hesitation towards the delivery of intervention components by women (i.e., helpline support, confirmatory testing, or community hub management) and would have preferred men delivering all intervention components.

Additionally, a few men and study implementers expressed mixed feelings towards the socio-cultural appropriateness of beerhalls as community hub locations during the prototype testing phase. While some men described beerhalls as open and safe spaces to discuss HIV prevention with male peers; others particularly with strong religious beliefs, feared being labelled as promiscuous or being stigmatised by their communities.

To mitigate structural and socio-cultural barriers, male participants and HCWs suggested confirmatory testing and PrEP initiation/distribution in communities rather than at clinics. While this adaptation went beyond the feasibility of this intervention, other recommendations were incorporated. For example, to increase helpline access, the helpline operating hours were expanded past men’s working hours until 7pm and on Sundays from 8:00am-3:00pm. The study team also employed both male and female helpline operators based on the participant’s gender preference. Lastly, although the sole use of beerhalls as community hubs was unintentional in this study and was selected based on availability-in the subsequent randomised controlled trial, various locations were selected as community hubs to encompass men’s preferences as well as to encourage secondary distributor enrolment and another wave of peer-delivered HIVST.

## 4. DISCUSSION

This IMEPRATIVE peer-delivered HIVST intervention was made more acceptable, feasible and potentially effective among men in eastern Zimbabwe through co-adaptation using a novel iterative prototyping approach prior to randomised controlled trial testing. Multiple qualitative methods and concurrent prototype testing enabled feedback across stakeholder groups and implementation levels. All stakeholder groups supported the intervention and anticipated positive effects on HIV testing and PrEP initiation among Zimbabwean men. Among men who tested the intervention, most reported oral HIVST use (primary distributors= 100%; peers recruited by primary distributors = 90%; peers recruited by secondary distributors = 63%) and approximately a quarter reported PrEP initiation (primary distributors= 25%; peers recruited by primary distributors = 30%; peers recruited by secondary distributors = 25%). Despite these encouraging findings, participants identified areas for refinement related to recruitment, information dissemination, incentive structure, and structural and socio-cultural factors which were either adapted and tested or may be considered for future interventions.

Our iterative prototyping approach helped us to understand how participants viewed peer-delivery across multiple levels - in terms of how men approached their own peers (e.g., interpersonal level) and how they conceptualised peer-delivered HIVST among men in the Zimbabwean context (e.g., community level) (56). Most participants described approaching or being approached by a male peer as appropriate and trustworthy. Men generally had a broad understanding of who they considered a “peer” and reported delivering the intervention to family and community members, suggesting that future research should further examine how Sub-Saharan African men conceptualise peers and friendship in relation to HIV interventions (30,31). Some participants also wished to extend HIVST distribution to male youth (<18 years), indicating a potential role for older peers and male role models, as demonstrated in Uganda, Zambia and South Africa (57–59). Others expressed interest in providing HIVST kits to female partners; although beyond the scope of this study, male-to-female test delivery remains under-researched and warrants further investigation (60).

In contrast to whom the participants found appropriate to approach, participants identified social groups which they considered culturally inappropriate to recruit – including older men, some occupational groups (e.g. farmers), and men with strong religious affiliations. We found that these social networks may have been underrepresented in this study. Consequently, the subsequent randomised controlled trial sought to recruit primary distributors across age, occupational, and religious groups to increase reach into diverse peer networks (61).

The co-adaptation process also identified important refinements in terms of information dissemination and incentive structures. Although men who tested the intervention were generally willing to use HIVST and initiate PrEP, this stood in contrast to reports from other men, CAB members, and study implementers who perceived low community awareness of these services. This may indicate that while the larger community has limited knowledge about HIV prevention strategies, when male- and client-centred information is disseminated, willingness to use these interventions may increase. Thus, based on participant feedback, we adapted the written materials to the needs of men. Information dissemination may be further supported through multi-media platforms, radio shows, film (62,63), social media campaigns (1,64,65), or at sporting events (1,66). For example, the EPIC-HIV application which aimed to provide information on HIV testing and linkage to care for South African men, was found to be an acceptable and appropriate method of information delivery (67). Additionally, men frequently relied on verbal instructions, demonstrations, and assistance from peers, family members, partners, or HCWs, indicating that social support extended beyond HIVST delivery. Future interventions could therefore consider assisted HIVST, peer accompaniment to clinics, or adherence support for PrEP and ART (23,68–70).

Furthermore, multiple stakeholders described areas for refinement surrounding the incentive structure. Stakeholders agreed that incentives increased motivation to deliver the intervention, although some requested higher compensation while others raised concerns about possible coercion. In this study, financial compensation was not increased because of budget constraints, yet men can distribute more kits during the larger trial and thereby receive more incentives. Future interventions may consider providing men and stakeholders with compensation through diverse delivery methods or at varying frequencies to maintain real-world feasibility while integrating a person-centred approach (71,72). Alternatively, different types of compensation including non-monetary incentives (i.e., food vouchers) as have been recently trialled among men in South Africa (73), may also be appropriate. The feasibility of interventions including incentive components also raises the importance of integrating interventions at scale through national programmes to ensure sustainable financing.

Finally, participants described some structural and socio-cultural factors, including long travel distances and limited clinic opening hours, which reduced (or could reduce) access to confirmatory testing and PrEP initiation. While we expanded the helpline operating hours to increase testing support for men, future interventions may consider differentiated service delivery (DSD) models to support access to HIV confirmatory testing and PrEP initiation outside of clinic settings. Effective DSD models may include peer-referral to: mobile clinics (65,66), pharmacies providing PrEP (74,75), or to PrEP e-delivery programmes (76) delivered by male stakeholders (65).

The refinements identified in this study were enabled by our iterative prototyping framework, which concurrently implemented multi-method qualitative research alongside prototype testing. This approach provided insight into intervention feasibility (per forum theatre workshops), contextual appropriateness (per the CAB meeting), group consensus about male social networks and intervention acceptability (per focus group discussions), individual experiences delivering or receiving the intervention (per in-depth interviews), and real-world implementation (observational reports). Whereas many public health interventions have been modified through sequential prototyping and pilot studies over extended periods (49,77); our approach provides an alternative strategy for low-resource settings or settings where interventions must be rapidly deployed.

While we took every precaution to implement the intervention effectively, this study was not without limitations. Each component of the draft intervention was evidence-based (32,78,79) and co-adapted via participant feedback; yet the intervention was not designed through a participatory research process in which community members define research questions, lead analysis, and disseminate findings (80). To address these limitations, the subsequent randomised controlled trial will employ peer researchers to co-lead data collection, analysis, and dissemination (36). We also recognize that our intervention may not have reached some men who could have benefitted from this intervention and were considered by primary distributors as culturally inappropriate to recruit (e.g., older men, religious men). Additionally, at the time of the study, two intervention components (the toll-free helpline number and the SMS-based HIV risk assessment tool) were not fully implemented and thus, perceptions on these components were often hypothetical in nature. Finally, while our results are promising and suggest that peer-delivered HIVST may be a valuable and effective intervention for Zimbabwean men, we urge our findings to be viewed with caution. In the second prototyping iteration, no peers recruited by primary distributors reported PrEP initiation compared with 25% in the first iteration despite multiple intervention adaptations, likely reflecting the small prototype sample size. Effectiveness will therefore need to be evaluated in the subsequent large-scale trial (36).

## 5. CONCLUSION

Through this study we have both co-adapted the IMPERATIVE trial’s peer-delivered complex intervention to more effectively support HIV testing and PrEP initiation among adult Zimbabwean men and developed a novel iterative prototyping framework combining a multi-method qualitative approach paired with prototype testing. Our co-adaptation process with multiple stakeholders allowed us to elicit areas for intervention refinement, namely to recruitment, information dissemination, the incentive structure, and to key structural and socio-cultural factors. The large-scale randomised controlled IMPERATIVE trial is now being conducted to test the co-adapted intervention and to determine its efficacy. If successful, mathematical modelling work will be done to assess its cost-effectiveness and sustainability at scale. Our iterative prototyping approach provides future study implementers (particularly in low-resource settings) across a wide range of health interventions with a tool to ensure intervention acceptability at all stages and to maximize a person-centred approach.

## Supporting information

Supplement 1: Qualitative data methodology

Supplement 2: Modified Information Sheet

## Acknowledgements

We would like to dedicate this research to Dr. Constance Nyamukapa who passed during the writing of this paper. Her endless dedication to global health and HIV prevention in Zimbabwe and her brilliant leadership will be deeply missed.

We would also like to acknowledge all research assistants for their assistance training, recruiting participants, transcribing data, and supporting data analysis. We would specifically like to thank Yeukai Bingepinge, Noster Chawira† and Tapiwa Magwenzi for their support and data collection through the helpline. We would also like to dedicate this research to Noster Chawira who passed during this study and was instrumental in project management. She is deeply missed by all.

We would like to acknowledge the village health care workers and guides who supported participant recruitment and would like to thank all participants who dedicated their time and expertise to this study.

## 6. DECLARATIONS

### Contributions

Qualitative interview guides were developed by authors RM, LC, MM, and MS. Qualitative data was collected by authors RM and LC, analysed rapidly by the entire qualitative team, and analysed in depth by author MM. Quantitative data was collected by the helpline team, including author FD and analyzed by author LM. Authors MM, RM, LV, LM, FD, PM, BT, PM, SG, CN, FT and MS contributed to the implementation of this intervention. These authors, in addition to WO, MI, GH, and TB, contributed to the wider study design. Author MM wrote the first draft of the manuscript and all authors read, provided feedback, and approved of the manuscript.

### Funding

This study was funded by the National Institute of Mental Health (R01MH133488, PI: FT). The funders had no role in the design of the study, data collection and analysis, decision to publish or preparation of the manuscript. Open access funding was enabled and organized by Heidelberg University and the Deutsche Forschungsgemeinschaft within the funding program Open Access Publikationskosten.

### Disclaimer

The findings and conclusions of this manuscript are those of the authors alone and do not necessarily represent the views of the National Institute of Mental Health.

### Conflict of Interest

Simon Gregson declares shareholdings in GlaxoSmithKline and Astra Zeneca. All other authors declare no competing interests.

### Participant consent for publication

Participants provided written informed consent for all intervention and study activities. No additional consent for publication was deemed necessary.

### Ethical Approval

This study obtained ethical approval from the Biomedical Research and Training Institute (BRTI) in Zimbabwe (AP187/2023) and the Medical Research Council of Zimbabwe (MRCZ; A/3098). We additionally obtained ethical clearance from the Stellenbosch University Medical Research Ethics Committee (MREC: M23/03/009) and the University of Lincoln in the United Kingdom (UoL2024_17988) and from Heidelberg University in Germany (S-232/2026). All research conforms with ethical principles stated by the Declaration of Helsinki.

### Provence and Peer Review

Not commissioned; externally reviewed.

### Data Availability Statement

The qualitative data and descriptive statistics used in this article are not publicly available but available upon reasonable request.

### Supplemental Material

Supplement 1: Qualitative data collection methods and qualitative data timeline

Supplement 2: Modified study information sheet

Supplement 3: Author reflexivity statement

### Open Access

This is an open access article distributed in accordance with the Creative Commons Attribution Non-commercial (CC BY-NC 4.0) license, which permits others to distribute, remix, adapt, build upon this work non-commercially, and license their derivative works on different terms, provided the original work is properly cited, appropriate credit is given, any changes made indicated, and the use is non-commercial.

## Notes

### Competing Interest Statement

One author, Simon Gregson declares shareholdings in GlaxoSmithKline and Astra Zeneca. All other authors declare no competing interests.

### Clinical Trial

NCT06370923

### Clinical Protocols

https://pubmed.ncbi.nlm.nih.gov/42281985/

### Author Declarations

The ethics committee of the Biomedical Research and Training Institute in Zimbabwe gave ethical approval for this work (AP187/2023). The Medical Resarch council of Zimbabwe gave ethical approval for this work (MRCZ: A/3098) The IRB of Stellenbosch University in South Africa gave approval for this work (MREC: M23/03/009). The IRB of the University of Lincoln in the UK gave approval for this work (UoL2024_17988). The IRB of Heidelberg University in Germany gave approval for this work (S-232/2026).

