## Supplement 1: Qualitative data methodology for "Engaging Zimbabwean men and stakeholders in the co-adaptation of peer-delivered HIV self-testing: iterative prototyping of the IMPERATIVE Trial"

### Supplement 1: Qualitative Data Collection

In this study, we utilised five distinct qualitative methods in parallel to iterative prototype testing and quantitative data collection. In this document, we expand upon manuscript section 2.4.2 describing the qualitative data collection procedures. We aim to present the details of our qualitative data collection to guide future replication of our study.

Our qualitative process included: i) forum theatre workshops, ii) a community advisory board meeting, iii) in-depth interviews, iv) focus group discussions, and v) observational reports implemented between November 2023 and May 2024. **Figure 2** further highlights the qualitative methodology timeline.

#### 1. Forum Theatre Workshops:

Three forum theatre workshops were conducted among men (n=12), healthcare workers (HCWs, n=12) and study implementers (n=11) in November 2023 prior to the first prototype testing iteration. Participants were specifically asked to i) provide insights on intervention feasibility and suggest recommendations to components they perceived as infeasible, ii) identify local resources to support implementation, and iii) discuss and adapt the intervention's *Theory of Change* framework (intervention inputs, processes, outputs, outcomes, and impact).

During the workshops, some study implementers (authors RM, FD, LC, TG, PM) acted out intervention components, such as HIVST kit use and peer HIVST delivery. Participants were encouraged to shout "freeze!" whenever they perceived a component as infeasible and to suggest or act out alternative approaches to advise the intervention.

#### 2. Community Advisory Board (CAB) Meeting:

Thereafter, in March 2024 a Community Advisory Board (CAB) meeting was held with 15 CAB members (2 / per IMPERATIVE trial site) to assess the cultural and contextual appropriateness of the intervention. The CAB specifically advised on i) study implementation, ii) the appropriateness of research and intervention methods and tools, iii) cluster randomization processes, iv) participant recruitment and retention methods, v) strategies for addressing study challenges, vi) anticipated community behaviours, and vii) result dissemination and policy development.

#### 3. In-depth Interviews (IDIs):

Additionally, we conducted informal in-depth interview (IDIs) with men (primary distributors=4; peers=11), HCWs (n=8), and community hub operators (n=2) who had tested the intervention prototype.

Men were asked about their experiences delivering or receiving the intervention; perceived intervention weaknesses; areas for intervention refinement; and recommendations to enhance the intervention. At the time of this iterative prototyping study, the toll-free helpline number and SMS-based HIV risk assessment tool were not fully implementable due (in part) to network connectivity challenges. Thus, participants were provided with airtime and private phone numbers of the study implementers to compensate for the helpline. Participants were asked about their anticipated perception of these intervention components.

Further, HCWs described their experiences providing HIV services to men linked to clinics through peer-delivered HIV self-testing and discussed acceptance of the SMS-based HIV risk assessment tool in clinics. Community hub operators reflected on their experiences distributing

HIVST kits within community hubs (i.e., beerhalls), including facilitators and barriers to distribution, storage conditions for HIVST kits, and experiences with incentive distribution.

**4. <u>Focus Group Discussions (FGDs):</u>**

These IDIs were supplemented by four focus group discussions (FGDs) conducted in May 2024 with men (n = 33, 8–9 men per FGD) who either did or did not test the intervention. These discussions explored group consensus around the acceptability and feasibility of intervention components, as well as perceptions of male peer social networks and broader facilitators and barriers surrounding HIVST and PrEP use. Prior to the FGDs, authors RM and LC facilitated an interactive drawing activity in which participants drew or listed the male peer groups they belonged to within their community (e.g., church groups, burial societies, beer-drinking clubs) and discussed the nature and frequency of interactions within these groups before proceeding to the FGD topic guide.

**5. <u>Observational Reports:</u>**

Finally, throughout the duration of the iterative prototyping testing (March-May 2024), eight observational reports were conducted where men and intervention stakeholders (including HCWs, community hub operators, and study implementers) were observed engaging with intervention components. For example, men were observed recruiting peers and delivering HIVST kits as well as engaging with confirmatory testing at local clinics. The aim of these observations was to better understand how men and stakeholders engaged with the intervention in a real-world setting.

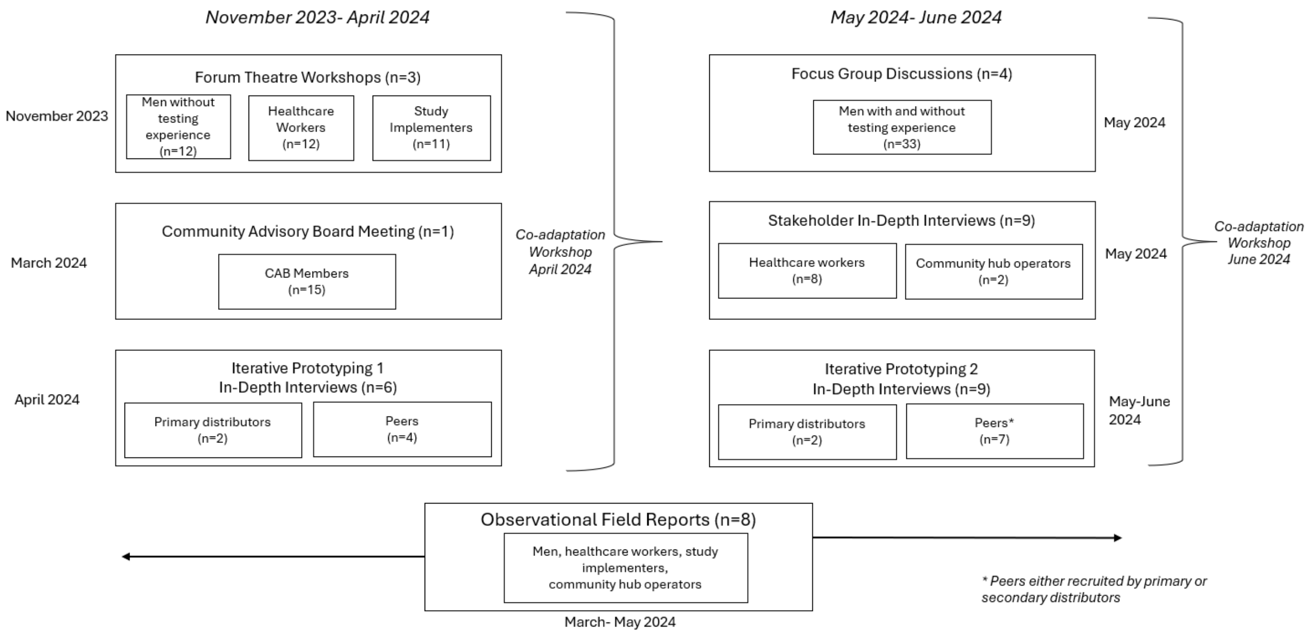


***Figure 2. Qualitative data collection timeline***
