## Supplement 2: Modified Information Sheet for "Engaging Zimbabwean men and stakeholders in the co-adaptation of peer-delivered HIV self-testing: iterative prototyping of the IMPERATIVE Trial"

**READ THIS INFORMATION LEAFLET FIRST, CONTACT THE FREE STUDY HELPLINE, AND READ A COPY OF THE CONSENT FORM THAT IS INCLUDED IN THIS PACKAGE**

**FREE STUDY HELPLINE NUMBER: 695**

#### PLEASE ATTEND ONE OF THESE CLINICS WHO ARE INVOLVED IN THE IMPERATIVE STUDY:

- ABERFOYLE CLINIC
- BONDA MISSION HOSPITAL
- EHPL 1 CLINIC
- EHPL 5 CLINIC
- EHPL 6 CLINIC
- GROOBI SPRINGS
- HOBHOUSE CLINIC
- LITTLE KRAAL PRISON CLINIC
- MAVHUDZI CLINIC
- MOYOWESHUMBA CLINIC
- MUTARAZI SATELLITE CLINIC
- MUTASA CLINIC
- NYANGA HOSPITAL
- NYAZURA CLINIC
- NYAZURA MISSION CLINIC
- OLD MUTARE HOSPITAL
- RUKWEZA CLINIC
- SADZIWA CLINIC
- SAGAMBE CLINIC
- SAKUPWANYA
- SAMANGA CLINIC
- SELBOURNE/NYANGA PINE CLINIC
- ST BARBRAS HOSPITAL
- ST JOSEPHS HOSPITAL
- TSONZO RURAL HOSPITAL
- ZINDI CLINIC

MANICALAND CENTRE
Public Health Research

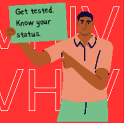


### IMPERATIVE study

- information sheet -

#### WHY ARE WE DOING THIS RESEARCH?

- Previous research has told us that men in Zimbabwe can find it hard to access HIV services including testing and treatment.
- This study will try to find ways to help men in Manicaland to overcome challenges accessing HIV testing, treatment, and prevention.

#### WHAT IS THE IMPERATIVE STUDY?

- This study is being run by the Manicaland Centre for Public Health Research.
- We want to find out if men in Manicaland have less barriers to accessing HIV services if men in the local community give out HIV self-test kits to their friends and other men in the same community

**READ THIS INFORMATION LEAFLET FIRST, CONTACT THE FREE STUDY HELPLINE, AND READ A COPY OF THE CONSENT FORM THAT IS INCLUDED IN THIS PACKAGE**

**FREE STUDY HELPLINE NUMBER: 695**

### WHAT DO YOU NEED TO DO?

**READ THIS SHEET FULLY AND FOLLOW THESE STEPS IN ORDER**

#### 1. CONTACT THE FREE STUDY HELPLINE ON 695

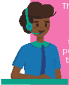

They will help you understand the study steps and support you in how to carry out your HIV self-test.

The study helpline team will also ask you to complete a short interview and check whether you are happy to take part in this study. A copy of the consent form is included in this pack. The helpline will try to contact you within 7 days if they have not spoken to you already.

***The helpline is free of charge and open every day (Monday to Saturday, 8:00 am to 7:00 pm and Sunday 9:00 am to 3:00 pm).***

#### 2. USE YOUR HIV SELF-TEST KIT

Which you have been given as part of this study. Read the information sheet and follow the instructions. HIV cannot be transmitted by saliva - the self-test detects antibodies.

**DO**

- Have a timer or clock available before you start.
- Follow all the instructions for using the kit as given in the information sheet.
- Tell the helpline if your kit is: damaged, lost or contaminated or past its use-by date and they will arrange for a replacement.

**DON'T**

- Do not use the kit if you are taking HIV treatment or using PrEP as the kit may give incorrect results.
- Do not eat, drink, or use mouth cleaning products (mouthwash, toothpaste or whitening strips) for 30 minutes before using the test.

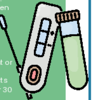

FREE STUDY HELPLINE NUMBER: 695

#### 3. DO NOT THROW AWAY THE TEST KIT

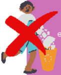

Put the kit in the disposal bag and place it in the brown envelope you received which also contains a form that will be used to help us record what happened after you took the test. Take these to the health facility with you.

#### 4. AFTER USING THE HIV SELF-TEST KIT, GO TO THE CLINIC

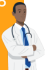

Go to a clinic as soon as possible to confirm the results. This will help them and you to determine whether you should start on HIV PrEP or HIV treatment. . You should visit one of the clinics listed on the back page of this leaflet.

***Remember to take your referral slip with you to the clinic***

#### 5. UNDERSTAND YOUR RISK OF HIV

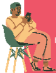

Learn about your risk of acquiring HIV using our free SMS tool. To use this tool, text **Start** to **695**. You will receive questions via SMS to help you understand if you may be at higher risk of acquiring HIV. The answers to these questions will not be recorded but can help you to understand if you would benefit from using HIV prevention. You can show the SMS to clinic staff who can advise you on PrEP.

#### 6. SMALL FINANCIAL REMUNERATION

The study helpline will call you to find out what happened and arrange to send you a small financial remuneration as a token of appreciation for your time. They will also ask if you would like to give our HIV self-test kits to your peers.

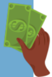
